# Keep Calm and Carry On: Projected Case Burden and Duration of the 2022 Monkeypox Outbreak in Non-endemic Countries

**DOI:** 10.1101/2022.05.28.22275721

**Authors:** Donal Bisanzio, Richard Reithinger

## Abstract

We report the estimated size and duration of a monkeypox outbreak in a simulated population of 50 million people with socio-economic -and demographic characteristics typical of a high-income, European country. Seeding an individual-based model with 3, 30, 300 primary cases, we estimate that—without public health emergency interventions—this could lead to a median of 18, 118, and 402 of additional secondary cases, respectively; similarly, the median duration of these outbreaks for these three scenarios would be 23, 37, and 37 weeks, respectively. We also estimate that contact tracing and isolation of symptomatic cases, or contact tracing, isolation of symptomatic cases and ring vaccination of the primary case contacts would substantially reduce the median size of outbreaks by 66.1—88.6% and median duration by 60.8–75.6%.

## Introduction

On May 7, 2022, monkeypox was detected in a traveler returning to the United Kingdom from Nigeria,^1^ and as of May 30, 2022, more than 400 additional confirmed cases were reported from North America, Europe, the Middle East and Australia.^2^ The disease is characterized by—at first—nonspecific symptoms and signs that include fever, chills, headaches, lymph node swellings, and myalgia, before smallpox-like rashes appear, which sequentially progress to macules, papules and vesicles.^3^ The disease is usually self-limiting, but may be severe in some individuals, such as children, pregnant women or the immunosuppressed; case fatality ranges between 1–10.6%,^4^ depending on the infecting virus clade. Although the natural reservoir of monkeypox remains unknown,^4^ rodents and non-human primates can harbor the virus; virus spillover is thought to occur through exposure to infected animal tissues and fluids (e.g., by eating undercooked bush meat). Human transmission of monkeypox virus is generally uncommon and results after close contact with lesions, body fluids, respiratory droplets, and contaminated materials (e.g., bedding, clothes). As per WHO^6^ monkeypox is endemic in 11 sub-Saharan African countries; cases reported outside of this geography are rare and associated with travel to endemic areas. Given the rapid increase and geographical expansion of cases in non-endemic areas since in early May, public health experts and the public have been on alert.^5^ On May 21, 2022, WHO convened a meeting of the Strategic and Technical Advisory Group on Infectious Hazards with Pandemic and Epidemic Potential (STAG-IH) meeting to discuss the current increase in monkeypox cases and whether it could pose a global health threat.

## Methodology

Using an individual-based mathematical modeling framework that has been applied to investigate the transmission of measles, Ebola, and SARS-CoV-2,^7^ we modeled a monkeypox outbreak in a simulated population of 50 million people with socio-economic -and demographic characteristics typical of a high-income, European country. Main features of the model were that it accounted for the high heterogeneity of people’s contacts and people’s mobility over short and long range in the simulated areas —key factors in the transmission dynamics and spread of infectious diseases. Outcomes of the model were median number of monkeypox cases, as well as the median duration of the outbreak. We modeled three baseline scenarios, with outbreaks seeded by the introduction of 3, 30 and 300 primary monkeypox cases in the simulated population. Baseline scenarios where no public health emergency interventions were performed were compared to two intervention scenarios, specifically (1) isolation of primary cases and contact tracing of individuals exposed to the primary case and isolating these in case of symptom onset, and (2) isolation of primary cases as well as contact tracing and vaccination of those exposed to primary cases (i.e., ring vaccination). A comprehensive model description, including model parameters used, is outlined in the Supplementary Material.

## Results

Our baseline scenarios project that—with no public health emergency interventions—the introduction of monkeypox infected individuals could lead to small national outbreaks of moderate duration; ultimately, the outbreaks would all subside. Thus, the model results estimated that the introduction of 3, 30, and 300 cases, without interventions, could cause a median 18, 118, and 402 secondary cases, respectively; similarly, the median duration of these outbreaks for the three scenarios would be 23, 37, and 37 weeks, respectively (Table 1). Conducting contact tracing with isolation of symptomatic cases would reduce the number of secondary cases by 72.2%, 66.1%, and 68.9%, respectively (Table 1). Adding ring vaccination to contact tracing would reduce the number of secondary cases by 77.7%, 78.8%, 88.6%, respectively (Table 1). The two intervention scenarios showed that adopting interventions targeting contacts of primary cases could reduce the median duration of monkeypox outbreaks between 60.8% to 75.6% (Table 1).

**Table 1.** Estimated median number of secondary cases and median duration of outbreak.

| Primary seed cases | Secondary cases (95% CI) |  |  |
| --- | --- | --- | --- |
|  | No interventions | Contact tracing | Contact tracing and ring vaccination |
| 3 cases | 18 (0–124) | 5 (0–16) | 4 (0–10) |
| 30 cases | 118 (43–609) | 40 (14–66) | 25 (6–41) |
| 300 cases | 402 (87–2092) | 125 (45–479) | 56 (4–259) |
|  | Outbreak duration (in weeks) (95% CI) |  |  |
| 3 cases | 23 (4–77) | 9 (3–20) | 7 (4–15) |
| 30 cases | 37 (20–99) | 14 (8–26) | 10 (6–15) |
| 300 cases | 37 (19–121) | 14 (10–26) | 9 (6–15) |

## Discussion

Our model results align with prior research on monkeypox outbreaks—whether in endemic or non-endemic countries—that demonstrated the low human-to-human transmissibility of the virus and its comparatively low potential to result in large-scale, heavy-burden outbreaks.^8-12^ Moreover, our results align with prior monkeypox transmission models;^13,14^ of note is, however, that these models did not account for the heterogeneity of interaction among individuals, people movement, treatment seeking behavior of the target population, not the performance of contact tracing and ring vaccination activities.

An unusual feature of the current outbreak is that a disproportionate number of confirmed cases were reported in men who have sex with men (MSM) (i.e., so far only <10 women seem to be among the more than 350 reported cases). Currently, there is no evidence that monkeypox is transmitted sexually; rather, it is more likely that cases were co-incidentally introduced into one or more MSM communities, with various individuals then subsequently exposed during mass gatherings through the close contact with lesions, body fluids, respiratory droplets, and contaminated materials.

In countries currently reporting monkeypox cases, our model shows how a strong public health response—specifically tracing and surveillance of contacts, isolation of symptomatic cases, as well as ring vaccination—would substantially reduce the number of secondary cases and the duration of outbreaks, potentially as much as by 88.6% and 75.6%, respectively.

We note several caveats of our analyses. First, we seeded our scenarios with a fixed number of cases, introduced once. We believe that this represents current observed events, whereby country outbreaks originated from isolated cases travelling back from endemic areas, or as result of super-spreader events such as mass social gatherings. Future tracking of the current outbreaks will have to determine how repeated introductions of primary index cases may affect the progression of outbreaks. Second, model parameters were based on prior outbreaks in endemic and non-endemic countries. At this time, there is no indication that the transmission dynamics of monkeypox have changed (e.g., sexual or aerosol transmission), nor that it includes an enzootic reservoir. Third, we assumed that individuals who have been vaccinated against smallpox are provided with 85% effective cross-immunity against monkeypox; we did not account for loss of immunity since time of vaccination. While there are reports that seem to indicate some loss of immunity,^14^ based on available data, all cases reported currently are in people <45–50 years old, i.e., an age group that would not have received the smallpox vaccine after routine vaccination for smallpox was discontinued following the eradication of the disease in 1980.^15^

In conclusion, our findings align with WHO’s assessment that the overall public health risk at a global level is currently “moderate”.^16^ Currently observed outbreaks in non-endemic countries should fairly rapidly be contained, particularly if primary cases and their contacts seek access to health care services, isolate if symptomatic, and potentially get vaccinated.

## Supporting information

Supplemental Technical Appendix

## Data Availability

All data produced in the present work are contained in the manuscript.

## Authors’ contribution

DB and RR conceived the research, analyzed, and interpreted the data, and drafted the manuscript.

---

**Figure 1.**
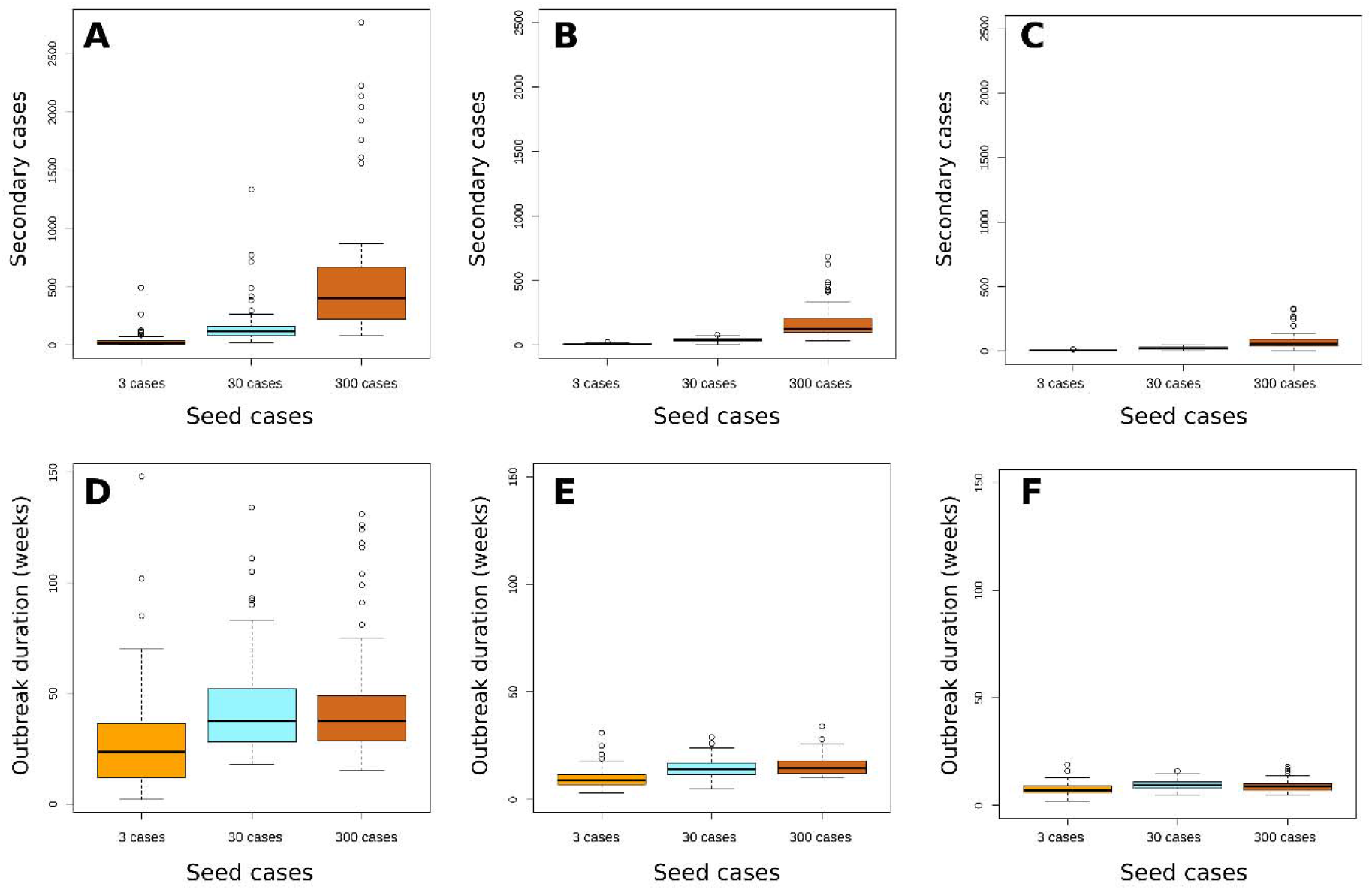
Median number of secondary cases and median duration (weeks) of monkeypox outbreaks using the MPX-IBM. Boxplots showing the median number of secondary monkeypox cases (Panels A – C) and duration of monkeypox outbreaks (Panel D – E) following seeding of 50 million in silico population with 3, 30, or 300 cases, comparing baseline scenario (Panels A and D) to intervention scenario 1 (Panels B and E) and intervention scenario 2 (Panel C and F). Solid lines, median; box, inter-quartile range; whiskers, 1.5 +/- inter-quartile range; o, outlier data points of the 1,000 simulations.

**Figure 2.**
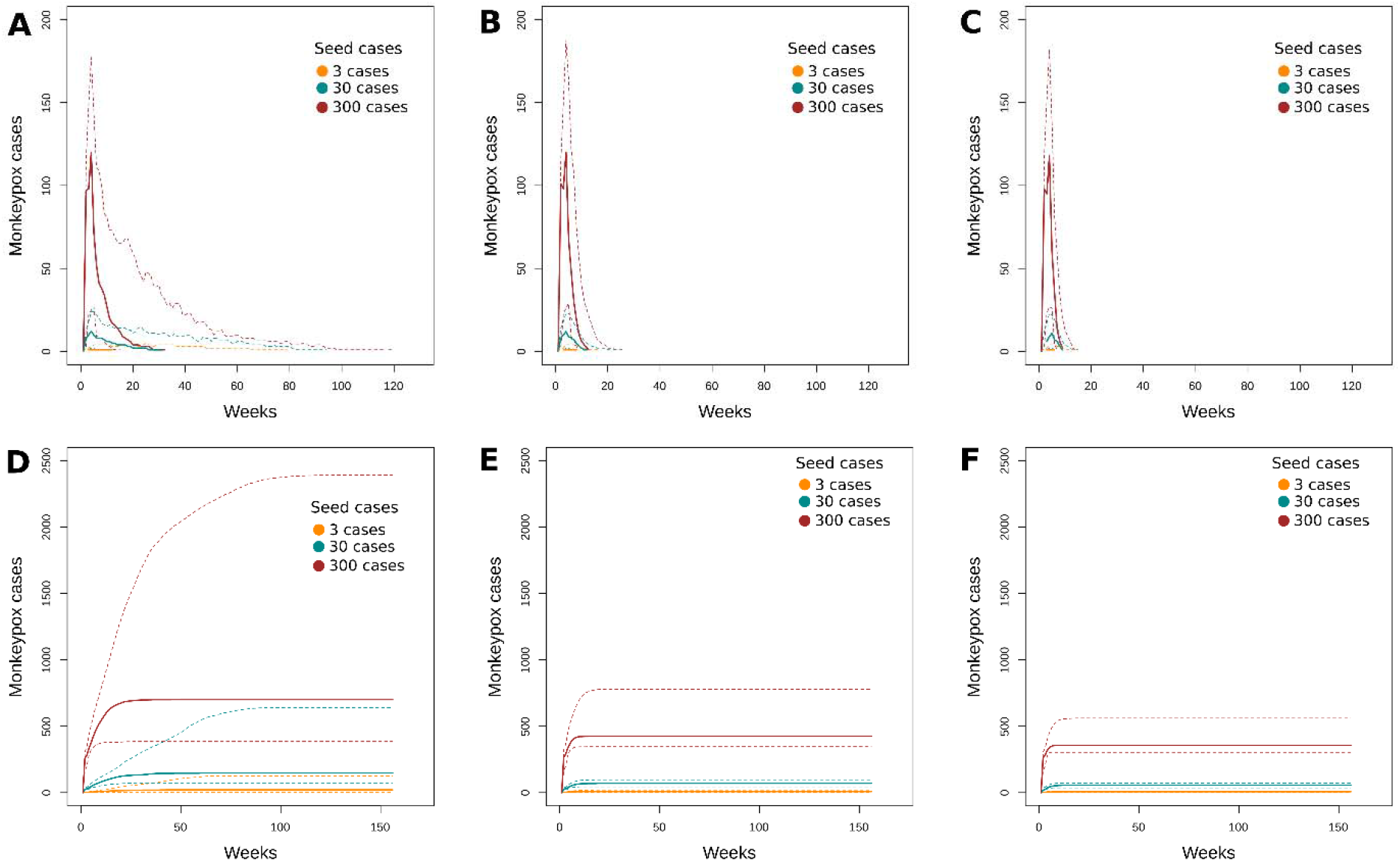
Epidemic curves and cumulative case count of simulated monkeypox outbreaks using the MPX-IBM. Panels A – C: Epidemic curves showing the number of primary and secondary monkeypox cases over time following seeding of 50 million in silico population with 3, 30, or 300 cases, comparing baseline scenario (Panel A) to intervention scenario 1 (Panel B) and intervention scenario 2 (Panel C). Panels D – F: Cumulative case count showing the number of primary and secondary monkeypox cases over time following seeding of 50 million in silico population with 3, 30, or 300 cases, comparing baseline scenario (Panel D) to intervention scenario 1 (Panel E) and intervention scenario 2 (Panel F). Broken lines: 95% confidence interval.

