## Supplemental Technical Appendix for "Keep Calm and Carry On: Projected Case Burden and Duration of the 2022 Monkeypox Outbreak in Non-endemic Countries"

**Data Dictionary**

*Demographic data.*

We modelled an outbreak of monkeypox using a simulated population of 50 million people, with characteristics of mean demographic statistics of the 27 European Union (EU) countries. The demographic statistics of the European Union were obtained from the Eurostat website (<https://ec.europa.eu/eurostat>). Demographic data we used to create the in-silico population included population age structure; average house size and house age composition; number of students per class; ratio of health workers to general population; enrolment of students for pre-school, primary school, secondary school, as well as university.

*Human Settlements.*

We disaggregated the 50 million population in 900 cells as a 30x30 matrix, representing settlements across 25 km^2^; the total landmass of the simulated country was equal to 562,500 km^2^, i.e., similar to the size of France and Spain. The population of each cell was estimated using a power distribution based on Zipf's law (Decker et al. 2007) fitted to a gridded population of the EU obtained from Eurostat (<https://ec.europa.eu/eurostat/web/gisco/geodata/reference-data/population-distribution-demography/geostat>).

**KSA-IBM Model Structure**

The following section provides a detailed description of the monkeypox individual-based model (MPX-IBM) human social network components. Given the transmission dynamics of monkeypox, incubation time, and disease duration lag, the model ran on a weekly time step. Weekly estimates made it easier to compare MPX-IBM estimates with reported outbreaks.

*Representing the Human Social Network (Intra-settlement Network).*

The MPX-IBM model structure accommodate the high heterogeneity of contacts among people—a key driver of infectious disease spread in communities. The MPX-IBM network was built to represent the “scale-free” and “small-world” characteristics common of human social networks. A scale-free network is a network characterized by a high fraction of nodes (individuals) connected to a low number of other nodes, while a few nodes have a high number of connections (i.e., so-called “super-spreaders”). The link distribution among nodes in the scale-free network can be described by a power-law distribution:

$$p\left( x \right)=x^{-\alpha}$$

where x is the number of links of a node, p(x) is the cumulative distribution, and the exponent α is the scaling factor. For human social networks, the power-law distributions have exponent α values ranging from 2 to 3 (Barrat et al., 2008). A network has a “small-world” attribute when two nodes in the network can reach each other through a short sequence of connected nodes (Kleinberg, 2002).

Interactions among individuals occurs in several locations, which can play a key role in the spread of disease agents. While people spend most of their daily time in their households, workplaces, and schools (Wang et al., 2015; González et al., 2008), interactions outside the routine locations (e.g., markets, shops, restaurants, and cinemas) are at the base of the small-world characteristic of human social networks. Additionally, the social network of an individual is linked to their age group, with people having more interaction with individuals of the same age group that they meet in different locations (Prem et al., 2017). Thus, for the MPX-IBM to accurately describe the interaction among people, we included in the model the distribution of interactions among individuals as well as the effect of individuals’ ages.

*Creation of contact network.*

After the population size and distribution of age groups of a settlement were estimated, a contact network among the simulated individuals was created using the following steps:

1. Households were created using the mean size of the family cluster. The number of people living together in each house was determined using a Poisson distribution, with the mean equal to the mean size of household inhabitants of EU countries (i.e., 2.4 people). The members of the household became the core contact network of each individual.
2. Links outside the household were calculated for each individual, based on a power-law distribution with α = 2.5.
3. The formula of a power-law distribution requires indicating a minimum value for *x*, with the minimum number of contacts for each individual equal to the number of their household members.
4. Each individual was linked to other individuals following contact matrices as the mean of the contact matrices of the 27 EU countries reported in Prem et al. (2017).
5. For all students attending pre-school, schools, and universities, a number of links extracted from a Poisson distribution with a mean equal to the average class size of EU countries were created; the links were connected with individuals of same age and from same settlement (square).
6. A location attribute was assigned to all links. We assigned four types: household, workplace, school or university, or other. The probability of assigning a link to a particular group was based on demographic characteristics of the EU population.

*Representing Human Movement Among Settlements*

The extra-cell spread of monkeypox in the MPX-IBM was represented with a weighted network that links settlements. The weight of each link was determined by the estimated mobility of people among settlements. The weight was calculated using a gravity model accounting for distance between settlements, and population size (Balcan et al., 2009; Kraemer et al., 2019). A gravity model is a modified law of gravitation that, in its most simple formulation (i.e., frictionless gravity model), considers the population size of two places and their distance apart to estimate the flow of people between them (Anderson, 2011). This method is not country-specific but has been used in the past in West Africa to model Ebola spread (Kraemer et al., 2019). The assumption of the gravity model is that larger settlements attract more people and settlements closer together share more people than distant ones. The gravity model created by merging population data and the distance matrix among the cells followed the methods described in Balcan et al. (2009) and Kraemer et al. (2019). The results of the gravity model was recorded in a flux matrix among settlements with columns and rows equal to the number of settlements. Each cell of the flux matrix contained the estimated population flow between two settlements.

**Modeling a Monkeypox Outbreak**

In the MPX-IBM, the infectious status of individuals followed the classical SEIR compartmental model structure: Susceptible (S) 🡺 Exposed (E) 🡺 Infectious (I) 🡺 Recovered (R). Infectious individuals were also at risk of being hospitalized. We did not include deaths in the model because the circulating monkeypox West African virus clade has not been reported as causing deaths outside of sub-Saharan Africa (CDC 2003). The transition from one status to another was a function of virus characteristics (e.g., virulence, incubation period, time to symptoms onset, and infectious period **Table S1**) and interaction among individuals (only for S to E). The MPX-IBM simulated the whole progression of the disease from exposure to skin lesions. We assumed that an infected individual will seek health services at onset of skin lesions. We started the simulation by seeding the in-sillico population with 3, 30, 300 cases. These seeding cases represent individuals that were exposed abroad and entered the country during their incubation period. We believe that the range of seeding cases we chose covers the number of cases that caused the outbreaks now reported across 21 countries in North America, South America, Europe, Middle East and Oceania, with the 30 and 300 seeding cases representing a super-spreading event.

**Table S1. Monkeypox transmission parameters of the MPX-IBM**

| Parameters | Value | Reference |
| --- | --- | --- |
| Transmission in household | 0.037 | Estimated from: Fine et al. 1988; Silenou et al. 2020; Usman et Adamu 2017 |
| Transmission outside household | 0.013 | Estimated from: Fine et al. 1988; Silenou et al. 2020; Usman et Adamu 2017 |
| Incubation period | 5–21 days (log normal distribution) | WHO 2022; CDC 2021 |
| Period flu-like symptoms | 1–5 days (uniform distribution) | WHO 2022; CDC 2021 |
| Rash and skin lesion | 14–21 days (log normal distribution) | WHO 2022; CDC 2021 |
| Infectious period | 14–28 days (uniform distribution) | WHO 2022; CDC 2021 |
| Probability to be symptomatic | 0.99 | Assumption based on WHO 2022 |
| Hospitalization probability | 0.90 | Assumption based on WHO 2022 |
| Time from symptom onset to hospitalization | 7–14 days (uniform distribution) | WHO 2022; CDC 2021 |

**Outbreak mitigation interventions**

The model included non-pharmaceutical and pharmaceutical interventions as implemented during the monkeypox outbreak that occurred in the US in 2003 (CDC 2003). The interventions were chosen as it was assumed that these interventions will be used by national health authorities in response to observed outbreaks, in accordance with current global guidelines (WHO 2022, CDC 2021). Non-pharmaceutical interventions (NPI) included the use of PPE for health care workers, contact tracing of exposed individuals, and increased treatment seeking in the population (both primary cases and the exposed) due to increased awareness (**Table S2**). The MPX-IBM took into account the marked spatial heterogeneity of contact tracing performance, by assigning each cell a contact tracing performance (i.e., contacts of symptomatic individuals successfully located by contact tracing teams) ranging between 20% and 90% (beta distribution). In the MPX-IBM, when a new case of monkeypox was identified, contacts of the infected individual were randomly selected using the cell contact-tracing performance and designated as the contact-traced group. Individuals in this group represent those exposed people who were monitored for 21 days and isolated if showing any symptoms. Thus, individuals in the contact-traced group had a low probability to infect others.

In the MPX-IBM’s second intervention scenario, ring vaccination was offered to contacts of a symptomatic individual. Acceptance of ring vaccination was set to 80% for all individuals in the contact-traced group, i.e., the mean vaccination coverage for two COVID-19 doses in EU countries (**Table S2**). Ring vaccination reduced an individual’s risk of becoming infected if exposed (**Table S2**), with ring vaccination targeting the contact-traced individuals completed within 2 weeks after the primary had been reported. Overall, NPIs and ring vaccination strategies were started in the MPX-IBM 1 week after the first cases of monkeypox were reported, similar to the approach that has been used during the current 2022 outbreak (WHO 2022).

**Simulations**

We tested 9 outbreak scenarios (3 scenarios with different number of seed cases x 3 intervention scenarios) withing the MPX-IBM, with 1,000 simulation runs performed for each scenario. The results of each scenario were median and 95% CI of secondary cases and outbreak duration (in weeks) calculated using 50%, 2.5%, and 97.5% percentile. The MPX-IBM was built using the Julia programming language (Bezanson et al. 2017) and the results were analyzed with the R language (R Core Team 2022).

Table S2. Response interventions

| Model input | Values/distribution | Reference |
| --- | --- | --- |
| Reduction in transmission probability due to use of PPE at health facilities during the post-intervention period | 70% | Assumed from Dunn et al. 2016; Jefferson et al. 2008; Kyaw et al. 2020 |
| Probability of seeking treatment at a health facility | 0.9 | CDC 2003 |
| Contacts reached by contact tracing system | 20–90% Uniform distribution | Assumption |
| Population > 45 years of age with smallpox immunity | 95% | Assumption |
| Smallpox vaccine efficacy against monkeypox | 85% | WHO 2022; CDC 2021 |
| Acceptance of vaccine during ring vaccination in future outbreaks, for all individuals in all rings | 80% | Assumed from WHO 2022b |

PPE = personal protection equipment
